# Prenatal Pyrethroid Exposure, Placental Gene Network Modules, and Neonatal Neurobehavior

**DOI:** 10.1101/2024.05.13.24307124

**Authors:** Yewei Wang, Jacqueline Holstein, Karen Hermetz, Amber Burt, Corina Lesseur, Parinya Panuwet, Nancy Fiedler, Tippawan Prapamontol, Panrapee Suttiwan, Pimjuta Nimmapirat, Supattra Sittiwang, Warangkana Naksen, Volha Yakimavets, Dana Boyd Barr, Ke Hao, Jia Chen, Carmen J. Marsit

## Abstract

Prenatal pesticide exposure may adversely affect child neurodevelopment which may partly arise from impairing the placenta’s vital role in fetal development. In a cohort of pregnant farmworkers from Thailand (N=248), we examined the links between urinary metabolites of pyrethroid pesticides during pregnancy, placental gene expression networks derived from transcriptome sequencing, and newborn neurobehavior assessed using the NICU Network Neurobehavioral Scales (NNNS) at 5 weeks of age. Focusing on the 21 gene network modules in the placenta identified by Weighted Gene Co-expression Network Analysis (WGCNA), our analysis revealed significant associations between metabolites and nine distinct modules, and between thirteen modules and NNNS, with eight modules showing overlap. Notably, stress was negatively associated with the interferon alpha response and Myc target modules, and the interferon alpha response module was correlated positively with attention, and negatively with arousal, and quality of movement. The analysis also highlighted the early and late trimesters as critical periods for the influence of exposures on placental function, with pyrethroid metabolites measured early in pregnancy significantly negatively associated with the protein secretion module, and those measured later in pregnancy negatively associated with modules related to oxidative phosphorylation (OXPHOS) and DNA repair. Additionally, the cumulative sum of 3-phenoxybenzoic acid across pregnancy was significantly negatively associated with the OXPHOS module. These findings suggest that prenatal exposure to pyrethroid pesticides may influence neonatal neurobehavior through specific placental mechanisms that impact gene expression of metabolic pathway, and these effects may be pregnancy period specific. These results offer valuable insights for future risk assessment and intervention strategies.

## Introduction

The pervasive use of pyrethroid insecticides in agriculture and residential areas has raised concerns regarding their potential neurotoxic effects in humans, particularly during vulnerable developmental stages such as gestation [1-5]. Pyrethroids, recognized for their efficacy in pest control, possess neurotoxic potential at elevated doses, warranting examination of their impact on fetal development [6-9]. The neurobehavioral effects are particularly alarming due to their implications for long-term health outcomes [6, 10, 11]. Although much of the evidence for neurotoxic effects on offspring originates from animal studies [6, 12, 13], a growing number of epidemiologic investigations also support these findings [14, 15]. Several studies have linked pyrethroids to behavioral disorders [16], cognitive deficits [17], learning disorders and attention deficit hyperactivity disorder [18, 19], further emphasizing the potential risks associated with these exposures.

Recent studies reveal a complex picture of the potential effects of prenatal exposure to pyrethroids on infants. Several studies document associations between exposure and compromised neural and mental development at one year of age, underscoring the need to minimize such exposures during pregnancy [21]. Paradoxically, other research suggests that exposure during the early and middle trimester might adversely affect motor and adaptive behavior in one-year-old children, although the underlying mechanisms remain elusive and could be influenced by confounding factors [22]. In contrast, certain studies report no significant effects on motor development at three months [7]. Another child cohort study suggests that pyrethroid exposure during gestation may not adversely impact early language development, potentially reducing the risk of low vocabulary scores [23]. These findings may indicate either positive effects or the influence of unmeasured confounders. The varied outcomes highlight the complexity of definitively linking prenatal pyrethroid exposure to specific neurodevelopmental trajectories and emphasize the need for further research to clarify these effects and inform public health guidelines.

The environmental health field increasingly recognizes gestation as a period of significant vulnerability and importance for human health [24, 25]. The placenta is a crucial organ during gestation, garnering considerable research interest for its roles in supporting fetal development and its potential influence on infant neurodevelopment and behavior [26-29]. Studies have indicated a notable link between various aspects of placental function—ranging from placental miRNA expression to genetic and epigenetic variation, including the expression of imprinted genes—and early neurobehavioral outcomes, as measured by the Neonatal Intensive Care Unit Network Neurobehavioral Scale (NNNS) [30-32]. These findings propose that the placenta could play a central role in shaping the early neurodevelopmental pathways of infants. Nonetheless, the research into prenatal exposures and their impact on fetal development must inherently consider placental function, given its mediatory role in fetal exposure to various substances [33-36]. Our recent investigations demonstrate the intricate interactions between environmental pyrethroids exposures and placental function, revealing these placental alterations may have significant implications for fetal development and other pathologic processes[37]. Still, significant gaps persist in understanding how these interactions influence NNNS outcomes in subsequent generations.

This research explores the associations of maternal urinary pyrethroid metabolite levels, placental functional gene networks, and neurobehavioral outcomes in neonates during critical developmental windows. We make use of the NNNS as an early comprehensive measure of developmental outcomes and examine the pyrethroid urinary metabolites levels across the pregnancy: 3-phenoxybenzoic acid (3-PBA), a principal metabolite of permethrin, cypermethrin, and deltamethrin, as well as 3-(2,2-dichlorovinyl)-2,2-dimethylcyclopropane carboxylic acid (both cis-DCCA and trans-DCCA metabolites), primarily derived from permethrin, cypermethrin, cyfluthrin and beta-cyfluthrin. By employing a comprehensive analytical framework, this study is designed to offer insights into the role of the placenta as a mechanism underlying the impact of prenatal pesticides on neurobehavioral outcomes in early life, thereby contributing to the burgeoning understanding of the placenta within the environmental health sciences.

## Methods

### 2.1 Study population

The SAWASDEE study aimed to examine the health of pregnant women involved in agricultural activities in Thailand and their newborn children. Recruitment took place between July 2017 and June 2019. Strict eligibility criteria were defined for participation, with detailed criteria available in Baumert et al. [38]. Participants were primarily recruited in their early trimester during antenatal care visits at district hospitals and community health clinics in Chiang Mai Province, Thailand. A total of 394 women were enrolled, 333 participants delivered live infants, and pyrethroid exposures were assessed in 332 of those women. Additionally, 253 participants had placenta samples available suitable for transcriptomic analysis. Based on health assessments, 320 of these infants, at 5 weeks of age, underwent the NNNS procedure at the SAWASDEE clinic. When paired datasets of infants and their mothers were considered, the total study sample was finalized at 248 pairs. All research protocols and procedures were rigorously reviewed and approved by the Institutional Review Board of Emory University and the Ethical Review Board of the Institute of Health Sciences, Chiang Mai University.

### 2.2 Urine collection, composition, and pyrethroid metabolites measurement

Urine samples were obtained up to 6 times during pregnancy at each antenatal care visit by first urinating briefly to waste, then collecting the remainder in a 500 mL cup. The samples were transferred into 2-3 3-oz. Qorpak bottles, labeled, and stored at -20°C. The time of collection, previous void time, and total volume were recorded. To provide trimester-specific exposure measures while keeping study costs reasonable, we created trimester-specific urine composites from each participant but retained the original discrete samples for later use. Detailed information on the urine composite procedure is available in Baumert et al. (2022) [38].

Prior to the analysis of urinary pyrethroid metabolites, all composited samples were randomized using the Fisher-Yates shuffle [39]. c-DCCA, t-DCCA, 3-PBA, were measured in urine using on-line solid phase extraction (on-line SPE) liquid chromatography (LC) coupled with negative mode electrospray ionization (ESI)-tandem mass spectrometry (MS/MS) with isotope dilution quantification. Before extraction, urine samples (200 ul) were enzymatically digested using purified β-glucuronidase and sulfatase enzymes (derived from H. pomatia) to deconjugate the target analytes. The enzymatically digested samples were centrifuged, transferred to autosampler vials, and injected onto a column switching system for extraction of the target analytes on a Strata RP on-line solid phase extraction (SPE) column. The on-line SPE column was washed with a mixture of acetonitrile: Milli-Q water (10:90, V/V) solution and the target analytes were eluted from the on-line SPE column to a Poroshell 120 EC-C18 analytical column for chromatographic separation. During mass spectrometric analysis, the target analytes were monitored using the multiple reaction monitoring mode. One quantitation ion and one confirmation ion were monitored for the native analytes, and one quantitation ion was monitored for the labeled analogues. Concentrations of the target analytes were determined using an equation derived from a matrix-matched standard calibration curve. In each analytical run, samples were analyzed concurrently with a 10-point calibration curve, one matrix blank sample, two matrix-matched quality control samples, and one solvent blank sample. The method demonstrated good precision. Quality assurance was maintained through semi-annual GEQUAS certification. To normalize pyrethroid metabolite concentrations, urinary creatinine was measured in each composited sample using LC-MS/MS [40].

### 2.3 Placenta collection and RNA extraction

Placental collection and RNA extraction were consistent with Baumert et al. (2022) [38]. Post-delivery, within a 2-hour window, five circular slices approximately 1 inch in diameter were excised from regions free of maternal decidua and around 2 cm from the umbilical cord insertion site. These sections underwent a rinse and were promptly immersed in RNALater solution (Invitrogen). After a minimum of 72 hours at 4°C, they were snap frozen using liquid nitrogen, and then homogenized into a powdered form. The freeze-dried samples were allocated into 2 mL cryovials and stored at -80°C until subsequent analysis. RNA extraction was performed using the Norgen Animal Tissue RNA Purification Kit. Concentrations of the resultant RNA were determined with a NanoDrop 2000 spectrophotometer, while RNA quality was ascertained through gel electrophoresis on an Agilent 2100 Bioanalyzer system.

### 2.4 Transcriptome analysis

Our approach to transcriptome analysis adhered to the method described by Li et al. (2023) [41]. Briefly, RNA-Seq libraries were prepared after rRNA depletion using the RNAse H protocol from New England Biolabs (NEB). Sequencing was carried out with DNBseq™ technology, targeting approximately 20 million paired-end 100 bp reads for each sample. Over 85% of these reads aligned to the human genome (GRCh38). Quality of the sequence reads was assessed using FastQC and MultiQC. Alignment and quantification were executed using STAR and the GENCODE v33 annotation, respectively [42]. Gene-level read counts were derived from FeatureCounts. Post filtering, which removed genes with low expression, genes from sex chromosomes, and non-protein-coding genes, normalization was executed using the DESeq2 R package [43]. The removeBatchEffect function from the limma R package addressed technical variances from RIN values [44]. After specific exclusions, the refined dataset incorporated 253 samples.

### 2.5 NICU Network Neurobehavioral Scale (NNNS) assessment

The NNNS, which can be employed at or near birth and during early infancy has also been demonstrated to be a useful tool for evaluating early developmental outcomes affected by prenatal exposures [9, 20]. At five weeks post-birth, mothers and infants visited the SAWASDEE clinics as detailed in Baumert et al [38]. Infants’ weight, length, and head circumference were documented, while mothers completed the Depression Anxiety Stress Scale (DASS). The NNNS, tailored for at-risk infants, was used to evaluate infants’ neurological functions and behaviors. Suitable for infants aged 30-46 weeks (adjusted for conceptional age), the NNNS is both internally and externally valid [38]. Study nurses, accredited by a Brown University trainer and unaware of infants’ exposure details, conducted the tests. These nurses participated in monthly reliability sessions. Assessments were conducted in a quiet, temperature-regulated room, starting with baseline observations, and progressing through habituation tests, reflex examinations, and other neurological evaluations. For the analysis, measurements associated with stress were aggregated under the “ Stress/abstinence “ category. The NNNS data were then organized into 13 distinct categories, including Arousal, Asymmetric Reflexes, Attention, Excitability, Handling, Hypertonicity, Hypotonicity, Lethargy, Non-optimal Reflexes, Quality of Movement, Self-regulation, and Stress/abstinence. Statistical summaries for each category were calculated, encompassing the mean, standard deviation (SD), minimum (Min), 25th percentile, median, 75th percentile, maximum (Max), and interquartile range (IQR), providing a comprehensive overview of the distribution and variability of each NNNS characteristic.

### 2.6 Association analysis between placental genes and NNNS

To examine gene-specific associations between transcript level and NNNS measurement, we used linear regression to investigate the associations between each gene counts with each NNNS measurements. Genes demonstrating significant associations at a nominal threshold (P < 0.05) were cataloged. The comprehensive gene list, alongside a frequency analysis of the genes correlated with NNNS, was visually presented. Bar charts illustrated the overall gene associations, and a heatmap provided a detailed visualization of the gene frequencies in relation to NNNS measurements.

To examine functional networks of co-regulated genes, we utilized the weighted gene co-expression network analysis (WGCNA), previously reported resulting in 21 gene modules [41]. The first principal component of the genes counts within each module is used to define a module eigengene, which was then examined for correlation with each NNNS measurement using the Pearson correlation, incorporating a false discovery rate (FDR) correction. A heatmap was constructed to visually represent these associations.

### 2.7 Association analysis between metabolite levels, gene modules, and NNNS

To elucidate the interconnections among exposures, modules, and outcomes within our study, we utilized a structured Sankey diagram, crafted with the plotly library in R. Within the diagram, nodes symbolize the exposures, modules, and outcomes, while the connecting links denote the correlations between them. For a focused trimester-based analysis, three composite variables, T1, T2, and T3, were formulated by grouping metabolites from the respective early, middle, and late pregnancy periods. Pearson correlation was employed to determine the correlations. We further applied a linear regression analysis to evaluate the relationships between two variables, such as exposure and placental gene module or placental gene module and NNNS, controlling for several confounding factors, including maternal birth weight, maternal height, maternal BMI at the first visit, maternal age, gestational age, geographic location, and infant sex. Statistical significance was determined using a threshold of p < 0.05.

## Results

### Characteristics of the Study Population and NNNS Outcomes at Five Weeks Postpartum

In this study, 248 pregnant women from the SAWASDEE cohort, with complete pyrethroid exposure data across three time periods during pregnancy and available placenta transcriptomics, had children who completed the NNNS at five weeks postpartum (Fig. 1). Demographics of the population are reported in Table 1. The study population comprised participants with a mean maternal age of 25.86 years (SD = 5.31) an average early pregnancy weight of 64.77 kg (SD = 10.75) and a height of 153.98 cm (SD = 6.06). The infants’ gestational age at birth was on average 269.56 days (SD = 7.90 days) or 38.5 weeks, ranging from 37.3 to 39.7 weeks. Infant demographics showed a balanced sex distribution, with males accounting for 50.8% of the sample. Neonatal metrics revealed an average birth weight of 3.01 kg (SD = 0.41), a body length of 48.37 cm (SD = 2.57), and a head circumference of 32.86 cm (SD = 1.93). Additionally, the average placental weight was recorded at 546.06 g (SD = 108.49). These parameters of this sub-cohort align closely with the physical characteristics reported in prior studies of the parent cohort [37, 41].

**Table 1.** Descriptive characteristics of the study population (N = 248), Study of Asian Women and their Offspring’s Development and Environmental Exposures, 2017–2019.

| Characteristics | n (%) / Mean ± SD |
| --- | --- |
| Maternal age (years) | 25.86 ± 5.31 |
| Maternal weight (kg) | 64.77 ± 10.75 |
| Maternal length (cm) | 153.98 ± 6.06 |
| Gestational age (days) | 269.56 ± 7.90 |
| Infant weight (kg) | 3.01 ± 0.41 |
| Infant length (cm) | 48.37 ± 2.57 |
| Infant head length (cm) | 32.86 ± 1.93 |
| Infant gender | 126 (50.8%) |
| Placenta weight (g) | 546.06 ± 108.49 |

**Figure 1.**
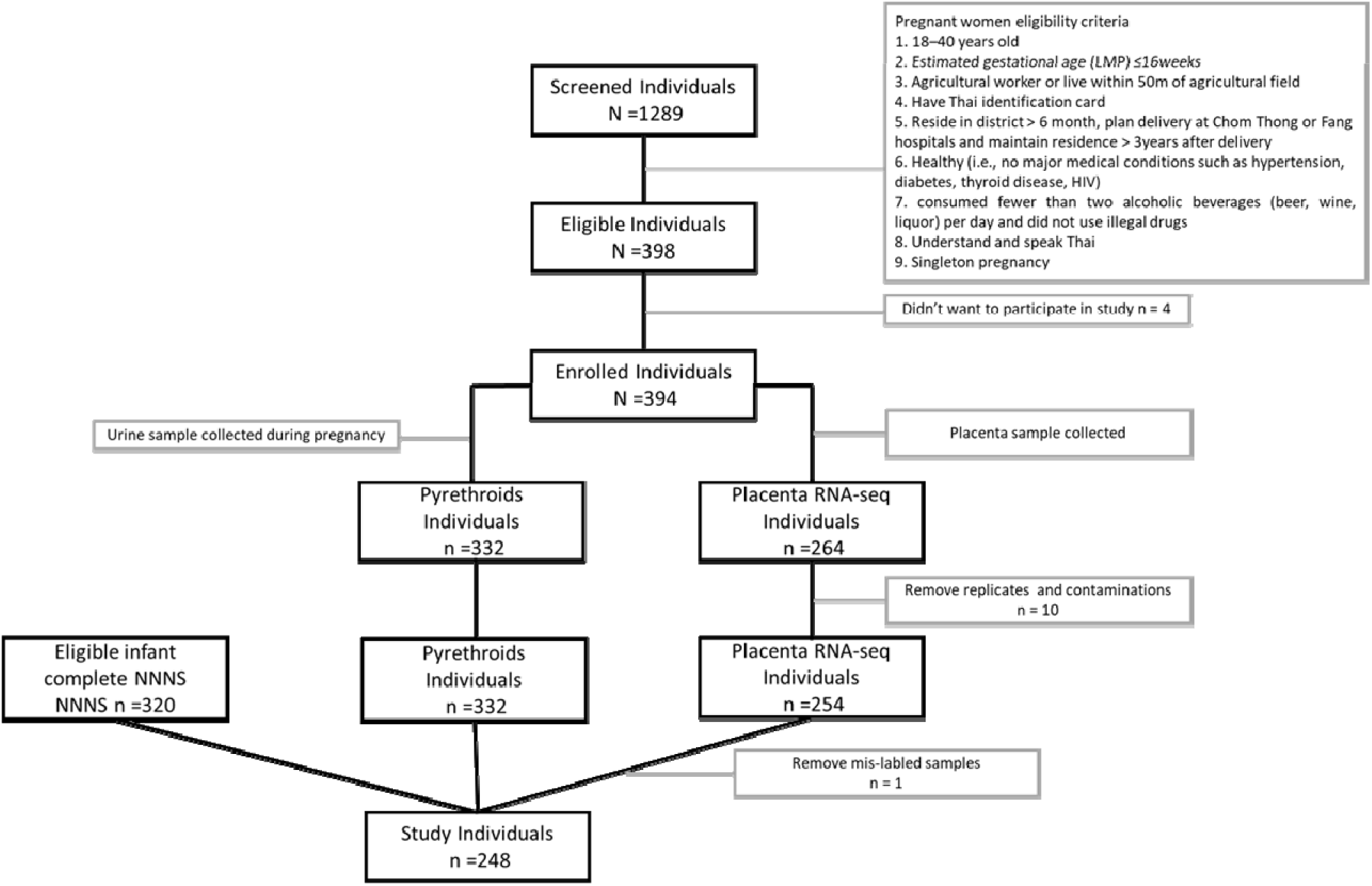
Participant selection in the SAWASDEE Study. This figure delineates the selection procedure for the study population in the SAWASDEE study, which investigates prenatal exposure to pyrethroid insecticides. Initially, 1289 pregnant individuals engaged in agricultural activities were identified within Chiang Mai Province, Thailand, between July 2017 and June 2019. Upon applying the inclusion criteria, 394 participants were selected for preliminary screening. Due to challenges in sample collection and quality, 62 individuals were excluded, resulting in 332 participants with pyrethroid exposure data. Additionally, 254 participants with placental RNA-seq data were identified. Finally, of the 320 eligible infants with NNNS data, 248 had complete data on maternal urinary pyrethroid metabolites and placental RNAseq.

Statistics for the NNNS summary scores, assessed at 5 weeks of age for 248 infants are presented in Table 2. Compared to the normal ranges for NNNS as outlined in Provenzi, et al. [45], most candidates fell within the normal range for the majority of NNNS measurements. Exceptions included Arousal, Attention, Excitability, Quality of Movement, and Stress/Abstinence, where some proportion of infants deviated from the normative values.

**Table 2.** Characteristics of NNNS Measurements.

|  | Mean | SD | Min | 25th | Median | 75th | Max | IQR | Count and Percentage Within Normal Range (n/%)* |
| --- | --- | --- | --- | --- | --- | --- | --- | --- | --- |
| **Arousal** | 3.69 | 0.54 | 2.71 | 3.29 | 3.57 | 4.14 | 5.29 | 0.86 | 135/54.4% |
| **Asymmetric Reflexes** | 0.47 | 0.71 | 0 | 0 | 0 | 1 | 4 | 1 | 245/98.8% |
| **Attention** | 5.71 | 0.76 | 2.14 | 5.29 | 5.71 | 6.14 | 7.29 | 0.86 | 196/79% |
| **Excitability** | 1.27 | 1.65 | 0 | 0 | 0 | 2 | 7 | 2 | 119/48% |
| **Handling** | 0.19 | 0.23 | 0 | 0 | 0.13 | 0.25 | 1 | 0.25 | 235/94.8% |
| **Hypertonicity** | 0.04 | 0.22 | 0 | 0 | 0 | 0 | 2 | 0 | 247/99.6% |
| **Hypotonicity** | 0.07 | 0.26 | 0 | 0 | 0 | 0 | 1 | 0 | 248/100% |
| **Lethargy** | 3.19 | 1.13 | 1 | 2 | 3 | 4 | 8 | 2 | 227/91.5% |
| **Non-Optimal Reflexes** | 4.27 | 1.35 | 0 | 3 | 4 | 5 | 8 | 2 | 243/98% |
| **Quality of Movement** | 5.52 | 0.23 | 4.6 | 5.5 | 5.5 | 5.67 | 6.2 | 0.17 | 20/8.1% |
| **Self-Regulation** | 6.07 | 0.59 | 4.58 | 5.66 | 6.15 | 6.54 | 7.33 | 0.88 | 211/85.1% |
| **Stress /Abstinence** | 0.56 | 0.36 | 0 | 0.28 | 0.52 | 0.8 | 1.74 | 0.52 | 194/78.2% |

\*Normal range was used from Provenzi's paper.

### Associations Between Placental Gene Expressions and NNNS Measurements

Across all associations between individual gene transcripts and NNNS, we identified a total of 153 unique placental genes (Fig 2A) demonstrating an association with an NNNS score (P<0.05). Notably, measurements pertaining to self-regulation (92 genes), arousal (84 genes), and attention (84 genes) emerged with the most associations with gene expression (Fig 2A). The heatmap in Figure 2B illuminates the frequency of associations between the top ten placental genes with the most significant associations with the NNNS domains. Ubiquitin Specific Peptidase 25 (USP25) emerged as having the most relationships, followed by Tudor Domain Containing 3 (TDRD3), Nemo Like Kinase (NLK), Tubulin Folding Cofactor C (TBCC), tRNA Methyltransferase 61A (TRMT61A), RFNG O-Fucosylpeptide 3-Beta-N-Acetylglucosaminyltransferase (RFNG), and Generic Kinase Activity 6 Precursor (GKA6P) (Fig 2B). These genes were primarily associated with self-regulation and arousal.

**Figure 2.**
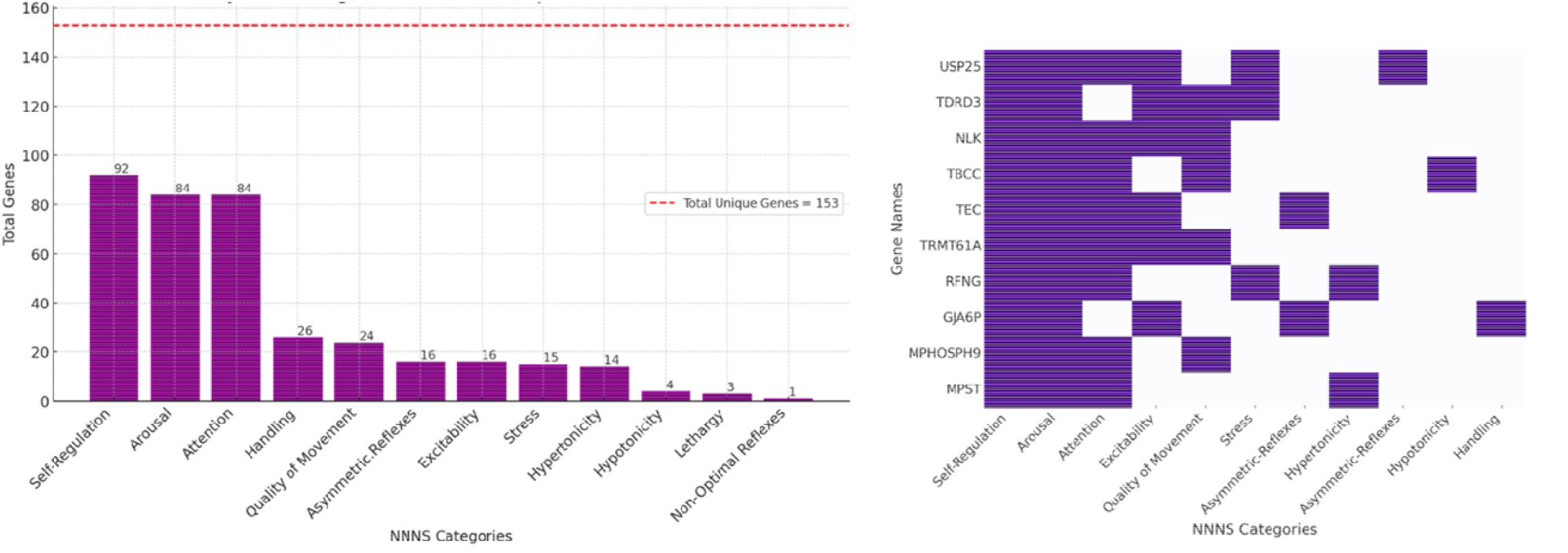
Significant Gene Associations with NNNS Measurements. This composite figure illustrates the association of significant genes with NNNS measurements. *Left Panel (A)*: A bar chart displaying the total number of specific genes associated with each NNNS measurement, with a total of 153 unique genes identified across all measurements. Each bar represents an NNNS measurement (x-axis) against the total number of associated genes (y-axis), with the exact number of genes denoted on each bar. *Right Panel (B)*: The heatmap showcases the associations of the top ten most frequently occurring genes with each NNNS measurement. The x-axis delineates the NNNS measurements, while the y-axis enumerates the names of these genes. Cells are shaded in dark purple when a gene is associated with the respective NNNS measurement, visually encapsulating the gene-neurobehavioral correlations.

### Associations Between Placental Gene Modules and NNNS Measurements

Pearson correlations were used to describe the relationships between placental gene modules, as previously delineated by WGCNA, and NNNS domains. Out of the 21 characterized placental gene modules, 12 modules showed significant correlations with NNNS domains (p<0.05) (Fig 3). Quality of movement exhibited the most correlations with modules, with eight, followed by attention and asymmetric reflexes, each with correlations to three modules, and Stress/abstinence with correlations to two modules. Although these NNNS domains demonstrated both positive and negative correlations with various modules, NNNS Stress/abstinence was distinctly negatively correlated with the interferon alpha response module and the Myc target module. The interferon alpha response module was the most frequently correlated module, with correlations to Attention, Arousal and Quality of Movement. This was followed by the unfolded protein response, DNA Repair and OXPHOS modules, each correlated with two measurements.

**Figure 3.**
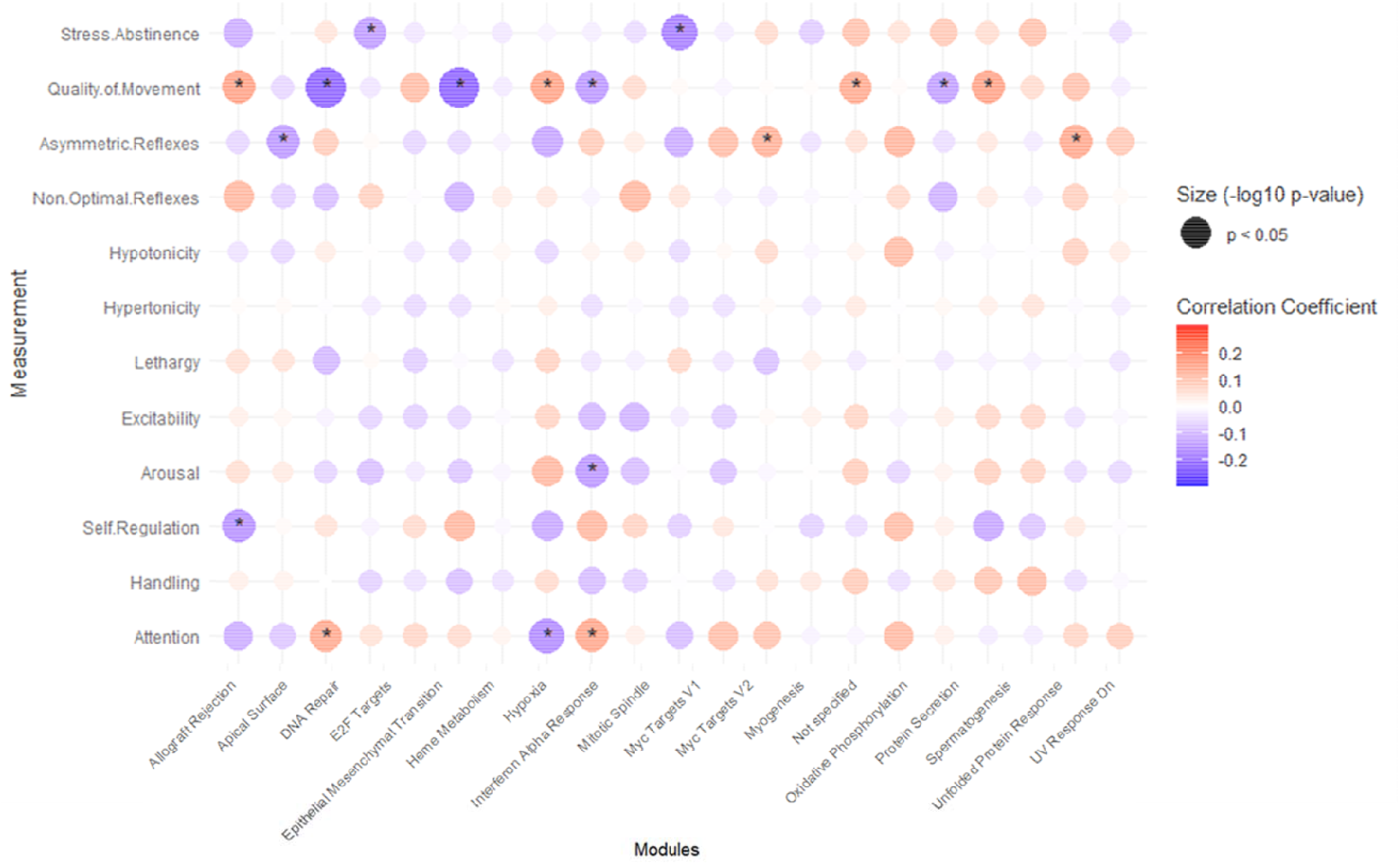
Correlations and Associations between NNNS and Gene Modules. A modified heatmap exhibits the coefficients of association between gene modules and NNNS measurements. The color intensity of each circle corresponds to the magnitude of the coefficient values, with darker shades indicating stronger associations. Red indicates positive associations and blue signifies negative associations. The size of the circles represents the -log10 p-value, which is inversely related to the p-values; thus, larger circles indicate lower p-values. Circles corresponding to statistically significant coefficients (p < 0.05) are marked with an asterisk (*).

### Associations Between Metabolite Levels, Gene Modules, and NNNS

A comprehensive Sankey diagram (Fig 4) visually depicts the complex interrelationships among metabolite levels, placental gene modules, and the NNNS using Pearson correlations to quantify the relationships. Notably, two pronounced significant negative correlations were observed within the -0.3 to -0.2 range: the mitotic spindle operations module and the DNA repair module, both correlated with movement quality. Nine distinct modules were significantly correlated with metabolites, while thirteen modules were significantly correlated to NNNS measurements, with eight overlapping modules identified. Of these eight modules, five showed consistent correlations: apical surface (positive), undefined (positive), protein secretion (negative), OXPHOS (negative), and spermatogenesis (negative). However, three modules—DNA repair, mitotic spindle, and OXPHOS—demonstrated complex associations. Furthermore, looking at correlations with metabolite data during specific periods of pregnancy, early (T1), middle (T2), and late (T3) trimesters, revealed that both the early and late trimesters demonstrated strong correlations between pyrethroid metabolites and placental modules. Metabolites in the early trimester were significantly negatively correlated with the protein secretion module, while third trimester metabolites showed significant negative correlations with modules related to OXPHOS and DNA repair. Notably, only the cumulative sum of 3-PBA across pregnancy revealed a significant negative correlation with OXPHOS module.

**Figure 4.**
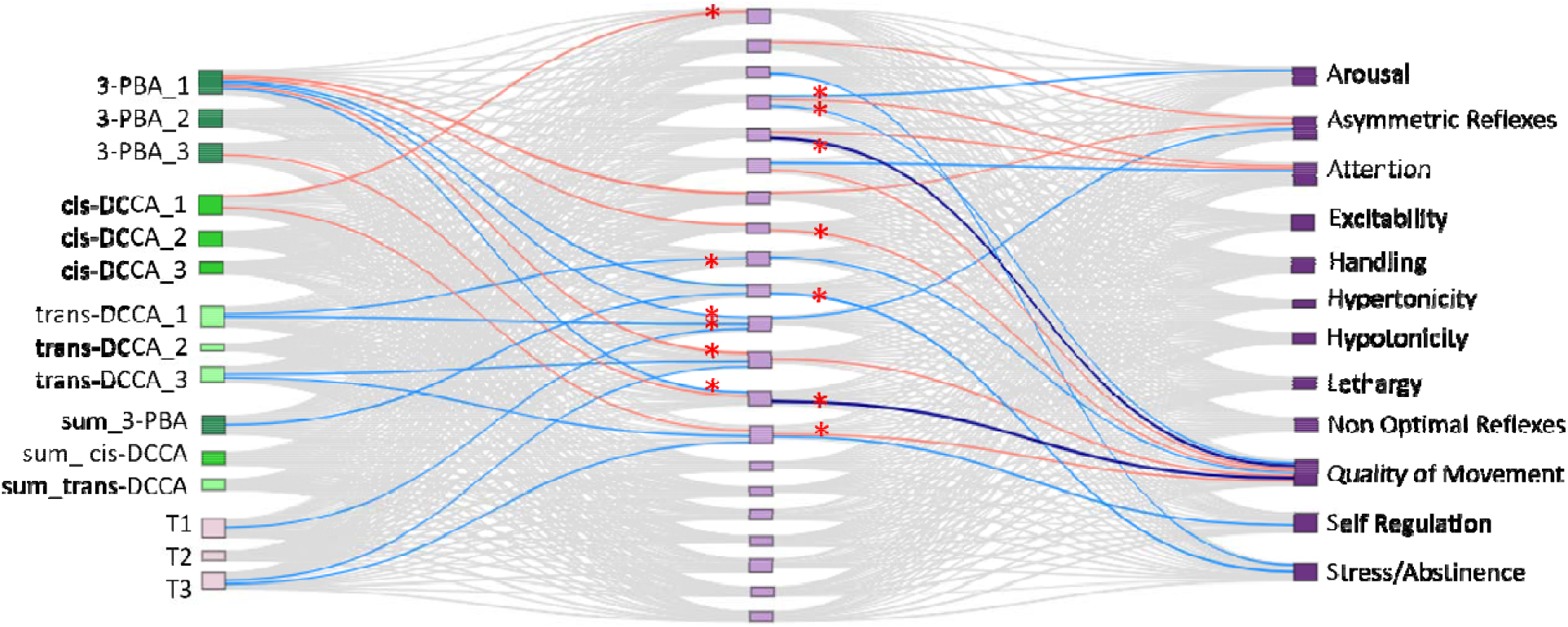
Sankey Diagram of Relationships Among Metabolites, Placental Gene Modules, and NNNS. This figure depicts a Sankey diagram elucidating the intricate relationships among metabolites, placental gene modules, and NNNS measurements. The relationships among these variables are assessed using Pearson correlation. Additionally, three derived variables, T1, T2, and T3, are computed by aggregating specific exposure columns within the same trimester, aiming to further illuminate the association between trimester windows and placental gene modules. The visual representation in the Sankey diagram employs color intensity and hue to signify the strength and direction of correlations for the flows between two modules. This figure illustrates correlation values and their associations using a color-coding scheme for distinct ranges: Significant positive correlations (0.1 to 0.2, p<0.05) are depicted in red; Significant negative correlations are differentiated by shade: light blue for -0.1 to -0.2 and dark blue for -0.2 to -0.3 (p<0.05); Negligible correlations (−0.1 to 0.1) and non-significance (p>0.05) are shown in grey. Color distinctions are also employed for various study components: metabolites are indicated by pink nodes; placental gene modules are represented by lavender nodes, and the NNNS node is marked with dark purple. The placental gene modules, displayed from top to bottom, are: *E2F Targets, Epithelial Mesenchymal Transition II, Myc Targets V1, Interferon Alpha Response, DNA Repair II, Unfolded Protein Response, Oxidative Phosphorylation, Spermatogenesis, Protein Secretion, Apical Surface, Not Specified, DNA Repair, Mitotic Spindle, Oxidative Phosphorylation II, Hypoxia, Myogenesis, Heme Metabolism, Epithelial Mesenchymal Transition, Allograft Rejection, Myc Targets V2, and UV Response Dn*. Red * indicates consistent significance based on the linear regression analysis, accounting for multiple confounding factors: maternal birth weight, maternal height, gestational age, maternal BMI at first visit, maternal age, geographic location, and infant sex.

Thirteen significant correlations identified in the Sankey diagram were confirmed through linear regression analysis, remaining robust even after adjusting for potential confounders (Fig 4, marked with *). Figure 5 illustrates the results of those linear regression models. 3-PBA in the first trimester demonstrated a significant positive association with the DNA Repair II module, suggesting that higher concentrations of this metabolite are associated with increased expression of DNA repair II module genes. cis-DCCA in the first trimester also showed a positive association with the DNA repair II moduleE2F Targets and Myc Targets V2 gene modules. Furthermore, significant associations were found between placental gene modules and NNNS outcomes. A negative association between the DNA repair module and the excitability outcome indicates that increased expression of the DNA repair module is linked to lower excitability scores in infants. Additionally, positive associations were observed between the mitotic spindle module and self-regulation, as well as between the OXPHOS II module and stress, suggesting that increased expression of these gene modules is correlated with higher scores in these NNNS domains.

**Figure 5.**
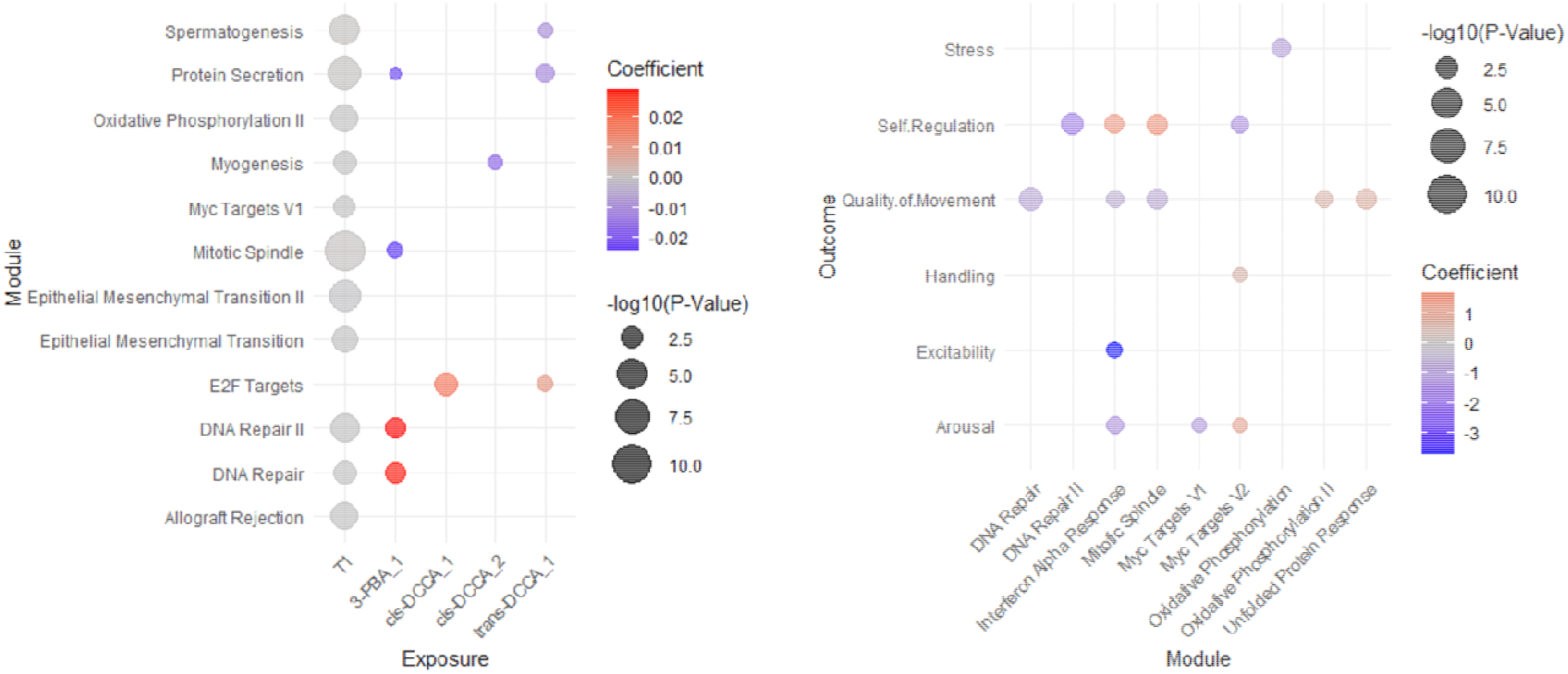
Bubble plots of Relationships Among Metabolites, Placental Gene Modules, and NNNS. This combined bubble plot illustrates significant associations across two domains: (1) the relationship between metabolites and placental gene modules, and (2) the relationship between placental gene modules and NNNS outcomes. The associations were analyzed using a linear regression model adjusted for confounders, including maternal birth weight, maternal height, gestational age, maternal BMI at the first visit, maternal age, geographic location, and infant sex. **Left Panel: Significant Associations Between Metabolites and Placental Gene Modules**. The x-axis represents the different metabolite variables, including 3-PBA (1, 2, 3), cis-DCCA (1, 2, 3), trans-DCCA (1, 2, 3). The y-axis represents biological modules derived from placental gene expression data (e.g., Spermatogenesis, Oxidative Phosphorylation II, DNA Repair, etc.). The coefficient represents the change in placental gene module expression (on the y-axis) for a 1-unit increase in metabolite concentration. The size of each bubble is proportional to the significance of the association (-log10(p-value)). Larger bubbles indicate more significant associations, with p-values below 0.05 indicating statistical significance. The color gradient from blue to red represents the direction and magnitude of the coefficient. Red indicates a negative association, while blue indicates a positive association. The intensity of the color reflects the strength of the coefficient. **Right Panel: Significant Associations Between Placental Gene Modules and NNNS Outcomes**. The x-axis represents the biological modules (e.g., DNA Repair, Mitotic Spindle, Myogenesis) that were found to be significantly associated with NNNS outcomes. The y-axis represents various NNNS outcomes, including Stress, Self-Regulation, Quality of Movement, Handling, Excitability, and Arousal. The coefficient represents the change in the NNNS outcome (on the y-axis) for a 1-unit increase in the expression of the placental gene module. The size represents the significance level (-log10(p-value)), with larger bubbles indicating stronger significance. The gradient from blue to red indicates the direction of the regression coefficient. Blue reflects positive associations, while red reflects negative associations, with more intense colors representing larger coefficient values.

## Discussion

Our study explores the associations of prenatal exposure to pyrethroids, quantified as maternal urinary metabolites (3-PBA, cis-DCCA, and trans-DCCA) levels during early, middle, and late pregnancy, on placental function and the subsequent associations with neonatal neurobehavioral development. We found significant correlations between pyrethroid metabolite levels and nine placental gene modules, as well as 13 modules correlated with NNNS measurements, including eight overlapping modules where five showed consistent correlations and three displayed complex interactions. Specifically, Stress/abstinence was negatively associated with modules related to the interferon alpha response and Myc targets, with the interferon alpha response module also significantly associated with attention, arousal, and quality of movement. Additionally, our analysis highlighted the early and late trimesters as critical periods for placental function, demonstrating significant negative associations of early pregnancy pyrethroid metabolite levels with protein secretion, later pregnancy metabolites with OXPHOS and DNA repair, and a persistent negative correlation of cumulative 3-PBA with OXPHOS throughout pregnancy.

Our study elucidates how the placenta, as a critical organ for fetal development, mediates the impact of pyrethroid metabolite exposure on fetal neurodevelopment health. Extant research highlights the placenta’s integral role in fetal neurodevelopment, particularly its capacity to modulate genetic responses to environmental and maternal stimuli that significantly shape the developmental trajectory of the fetus. Changes in placental gene expression profoundly affect infant neurobehavior, affirming the placenta’s role as a vital intermediary in neurodevelopment [28]. Additionally, structural changes and inflammatory responses within the placenta contribute to adverse developmental outcomes, reinforcing its critical position in developmental processes [46]. Studies also expose the placenta’s susceptibility to endocrine-disrupting chemical exposures, which compromise its endocrine functionality and immune interactions, potentially leading to neurodevelopmental disorders [47]. Furthermore, variations in CpG methylation within the placenta associated with cognitive functions in children born prematurely support the hypothesis that early placental adaptations can have enduring effects on child health [48]. Our findings build upon this framework by demonstrating that pyrethroid metabolite levels are related with differences in pivotal placental functions—specifically protein secretion, OXPHOS, and DNA repair—through co-regulated gene signatures. These pathways are critical during pregnancy, influencing conditions such as preeclampsia and fetal growth restriction, which are characterized by substantial oxidative damage and impaired repair mechanisms. Additionally, external factors like maternal smoking intensify the adverse effects of oxidative stress on both placental and fetal health, underscoring the crucial role of oxidative stress and mitochondrial dysfunction within the placenta across a spectrum of pregnancy outcomes [49, 50].

Our analysis also identified 153 unique placental genes significantly associated with NNNS measures, particularly affecting self-regulation, arousal, and attention. Notably, genes such as Ubiquitin Specific Peptidase 25 (USP25), which are linked to the migration and invasion of trophoblast cells as well as neurodegeneration, highlight potential areas for further investigation [49, 50]. These findings reinforce the importance of the placenta in fetal neurodevelopment and suggest that understanding the multifaceted roles of placental genes could provide valuable insights into mitigating adverse prenatal exposures.

Our findings also introduce a critical perspective on the temporal dimension of prenatal pyrethroid exposures and their relationship to placental functional pathways, highlighting a potential elevated susceptibility during the early and late trimesters, especially the first trimester. During these critical periods, our analysis revealed significant adverse correlations between the concentrations of pyrethroid metabolites and differences in placental gene modules. Specifically, the early trimester showed strong negative correlations between summed metabolite levels and protein secretion module, while in the late trimester, metabolites negatively affected modules associated with OXPHOS and DNA repair. Although 3-PBA levels showed no significant correlations with NNNS, we observed a consistent negative correlation between the sum of 3-PBA across all trimester and placental OXPHOS. These patterns highlight the placenta’s complex role in mediating the impact of environmental pyrethroid exposures on fetal neurodevelopment, which varies throughout different stages of pregnancy. Despite some inconsistent findings in the literature, the majority of research, including ours, supports a significant correlation between prenatal pyrethroid exposure and developmental effects, particularly during sensitive periods of gestation, with variability in effects across different pregnancy stages [7, 51-54]. For instance, elevated maternal urinary concentrations of 3-PBA during the third trimester have been linked to reduced mental development index scores at 24 months [51], underscoring the importance of later gestational exposure. Conversely, significant exposure during the early trimester has been associated with decreased motor and adaptive behavior scores in 2-year-old children [53], affirming its criticality in early fetal development. Further supporting this, increased risk ratios for autism spectrum disorders with higher second-trimester 3-PBA concentrations [54], and significant inverse relationships between 3-PBA levels and serum free triiodothyronine (FT3) in pregnant women, suggest potential disruptions in thyroid function [55]. These diverse outcomes underscore the complex interplay between pyrethroid exposure and neonatal thyroid hormone status and development. Building on these observations, our study not only reaffirms the importance of timing in pyrethroid exposure but also highlights the placenta’s pivotal role in mediating these effects, contributing to the growing body of evidence supporting the critical windows identified in epidemiological studies. This emphasizes the need for ongoing research into the specific mechanisms through which pyrethroids influence fetal development and suggests further exploration into protective strategies during vulnerable periods.

Despite being one of the largest of its kind to examine the relationship between placenta transcriptome and pyrethroid exposures, our study has several limitations. For example, given the extensive number of analyses performed, our sample size may provide an inadequate statistical power. Furthermore, while our results indicate that metabolite levels like cis-DCCA may be predictors of neurodevelopmental outcomes, the specificity with which these metabolites reflect actual neurobehavioral impacts requires additional investigation with longer follow-up. The placenta is a heterogeneous organ consisting of a variety of cell types, and our use of bulk transcriptomics does not account for cell-specific effects. Additionally, the homogeneity of our study cohort and the lack of covariate adjustment in our regression models may restrict the generalizability of our findings and impact the precision of our effect estimates. Importantly, our goal here was not to demonstrate causality, but instead to describe inter-relationships of pyrethroid metabolite levels, placental functional impacts, and early neurobehavioral performance, to inform specific hypotheses which can be assessed in a causal framework. Our study suggests that prenatal exposure to pyrethroid pesticides can alter placental gene expression of metabolic pathways, influencing neonatal neurobehavior. These insights lay a foundation for future research into the causal mechanisms underlying these effects.

## Conclusion

This study illuminates the comprehensive links between prenatal pyrethroid pesticide exposure, placental gene expression, and neonatal neurobehavioral development. It uncovers the complex interplay between environmental exposures and placental functional outputs at pivotal stages of neurodevelopment. These findings underscore the critical need for monitoring environmental pyrethroid exposures during pregnancy and demand a deeper exploration of how these exposures, in conjunction with other factors, shape the neurodevelopmental pathway from the earliest stages of life. Consequently, this work calls for a comprehensive research approach that includes environmental interventions and a focus on placental health to protect fetal development amidst growing concerns over environmental pyrethroid exposures.

## Data Availability

All data produced in the present study are available upon reasonable request to the authors

